# Epidemiology of cruciate ligament surgery in Japan: A retrospective cohort study from 2014 to 2019

**DOI:** 10.1101/2023.07.06.23292305

**Authors:** Shota Uchino, Masataka Taguri

**Affiliations:** Department of Data Science, Graduate School of Data Science, Yokohama City University, Yokohama, Japan; REHASAKU Co., Ltd, Tokyo, Japan; Department of Health Data Science, Tokyo Medical University, Tokyo, Japan

## Abstract

Understanding the incidence and trends of cruciate ligament (CL) surgeries in Japan is crucial for providing effective healthcare services. This study aimed to use open data available from the National Database of Health Insurance Claims and Specific Health Checkups of Japan (NDB) to determine the changes over time in CL surgeries and to analyze the characteristics of the Japanese population in terms of sex and age. We retrospectively identified CL surgeries of the knee joint registered from April 2014 to March 2020 using the NDB open data. Data on sex, age, and practice were extracted to determine the number of cases per 100,000 population. Trends in the annual incidence of CL surgeries were evaluated using Poisson regression analysis. In total, 112,686 CL surgeries were performed from 2014 to 2019. Arthroscopic ligament reconstruction accounted for 98% of all the cases. The number of surgeries performed had increased significantly from 16,975 in 2014 to 19,735 in 2019 (P<0.001). Overall, CL surgeries were most common in the 15–19 years age group, followed by the 20–29 years age group. However, these trends differed between males and females. The incidence of CL surgery in Japan has increased, and its characteristics vary by gender and age. These characteristics have been identified not only in younger patients but also in middle-aged and older patients. It would be valuable to further investigate the general patterns of CL surgery in Japan.

## Introduction

In recent years, the availability of medical and national big data has made it possible to determine the incidence of ligament injuries and surgeries [1–9]. These studies have revealed epidemiological models that allow for the generalization of disease incidence. They have also been used to improve treatment strategies and health care services to enhance patients’ quality of life.

In Japan, an epidemiological study using insurance records of middle and high school athletes reported 30,458 anterior cruciate ligament (ACL) injuries (0.81 per 1000 athlete-years) in these growing athletes over a 10-years from 2005 to 2014. Furthermore, the incidence rate of ACL injuries was reported to be 2.8 times higher in female athletes than in male athletes [10]. This previous study determined the incidence of ACL injuries using a large and long-term insurance dataset; however, it only focused on populations considered at high risk for ACL injury, and the incidence and the post-injury response of other populations remain unknown.

To understand the incidence and trends of the disease in the Japanese population, a larger sample size should be examined. Epidemiological studies using the National Database of Health Insurance Claims and Specific Health Checkups of Japan (NDB) established by the Ministry of Health, Labour and Welfare (MHLW) and NDB open data, which is a more generalized compilation of the NDB, have been actively conducted in the field of locomotor system [11–13]. Epidemiological modeling from these big data could lead to improved healthcare services.

This study aimed to elucidate trends in the number of cruciate ligament (CL) surgeries, sex, age distribution, and characteristics over time using NDB open data from 2014 to 2019. We hypothesized that the number of CL surgeries would increase over the years, and that there would be a higher incidence of such surgeries among females.

## Materials and methods

This population-based retrospective cohort study uses the NDB open data provided by the MHLW from April 2014 to March 2020. These data are widely accessed for research purposes and have been anonymized. Therefore, obtaining informed consent was not necessary.

We accessed the NDB open data website and obtained data on the "Number of calculations by sex, and age group" for "operation (Code K)" [14–19]. There were four categories of the CL of the knee joint: ligament tear suture (K074), arthroscopic ligament tear suture (K074-2), ligament reconstruction (K079), and arthroscopic ligament reconstruction (K079-2). Data on the number of registered CL surgeries were stratified by fiscal year, sex, and age (5-year interval). No detailed analyses were performed if the number of registered CL surgeries had no specific values shown in <10 cases. The sex and age data were obtained by calculating the number of surgeries per 100,000 population. The calculation method utilized population estimates published by the Statistics Bureau of the Ministry of Internal Affairs and Communications, based on population data as of October 1 of each year [20].

Changes in the number of surgeries on the CL of the knee joint over time were assessed using Poisson regression, with the number of surgeries as the outcome variable and the year of surgery as the explanatory variable. The supporting information file contains the details of the STROBE checklist (S1 Table). The NDB open data is widely available for research purposes by the Ministry of Health, Labour, and Welfare (MHLW). The data are compiled and published based on existing anonymized NDB, and since there is no corresponding table, individuals cannot be identified. Therefore, ethical approval was not required for this study.

## Results

In total, 112,686 CL surgeries were performed from April 2014 to March 2020, with 109,925 (98%) being arthroscopic ligament reconstruction. The remaining procedures included ligament reconstruction and arthroscopic ligament tear suture, each accounting for approximately 1.0% of the total number of surgeries performed, with very few cases of ligament tear suture (Table 1). The number of arthroscopic ligament reconstruction increased significantly from 16,997 (18.3 cases/100,000 population) in 2014 to 19,774 (22.1 cases/100,000 population) in 2019, surpassing the number of all other techniques (P<0.001) (Fig 1).

**Fig 1.**
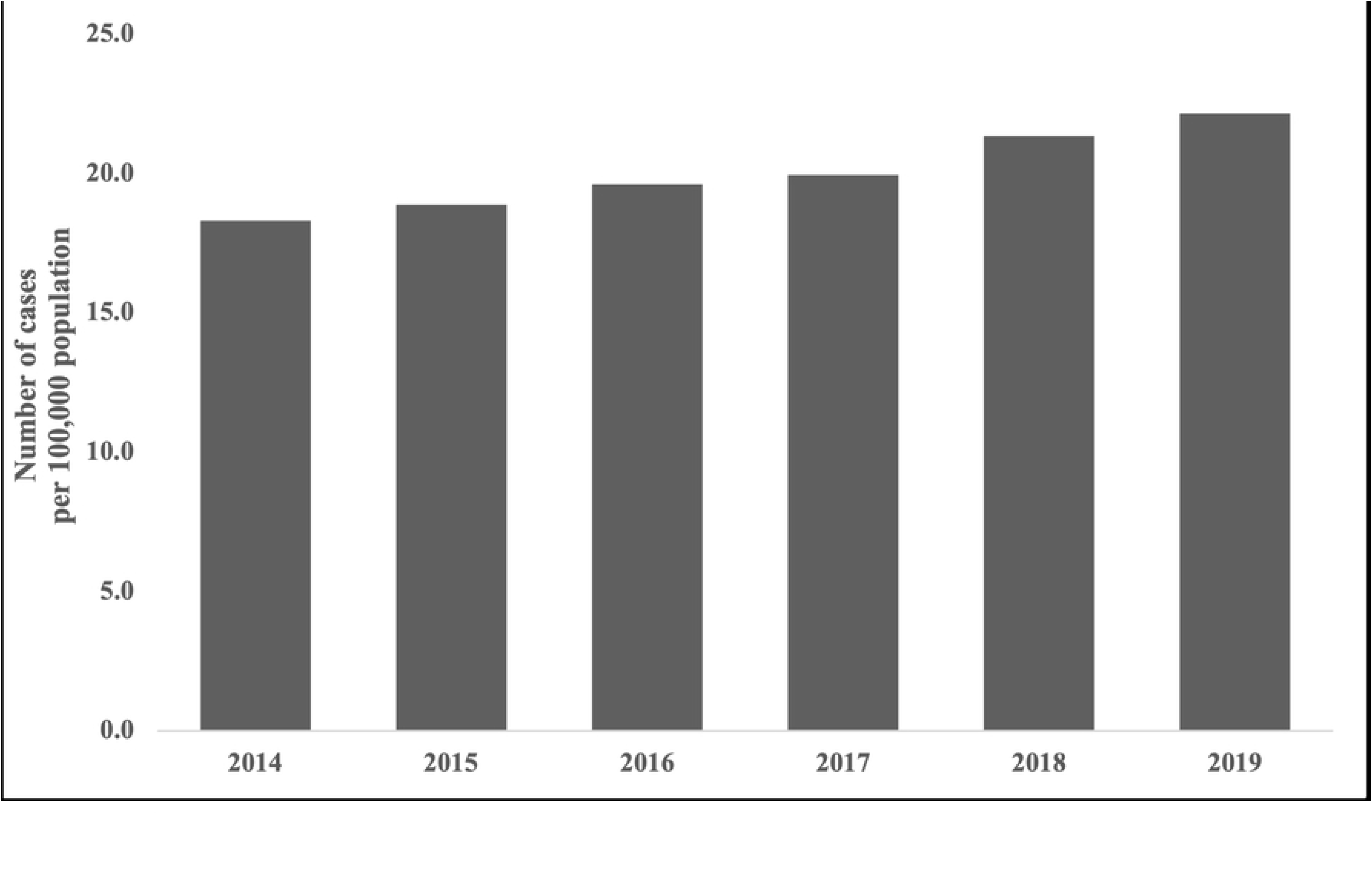
Number of registered arthroscopic ligament reconstruction procedures per 100,000 population from 2014 to 2019.

**Table 1.**
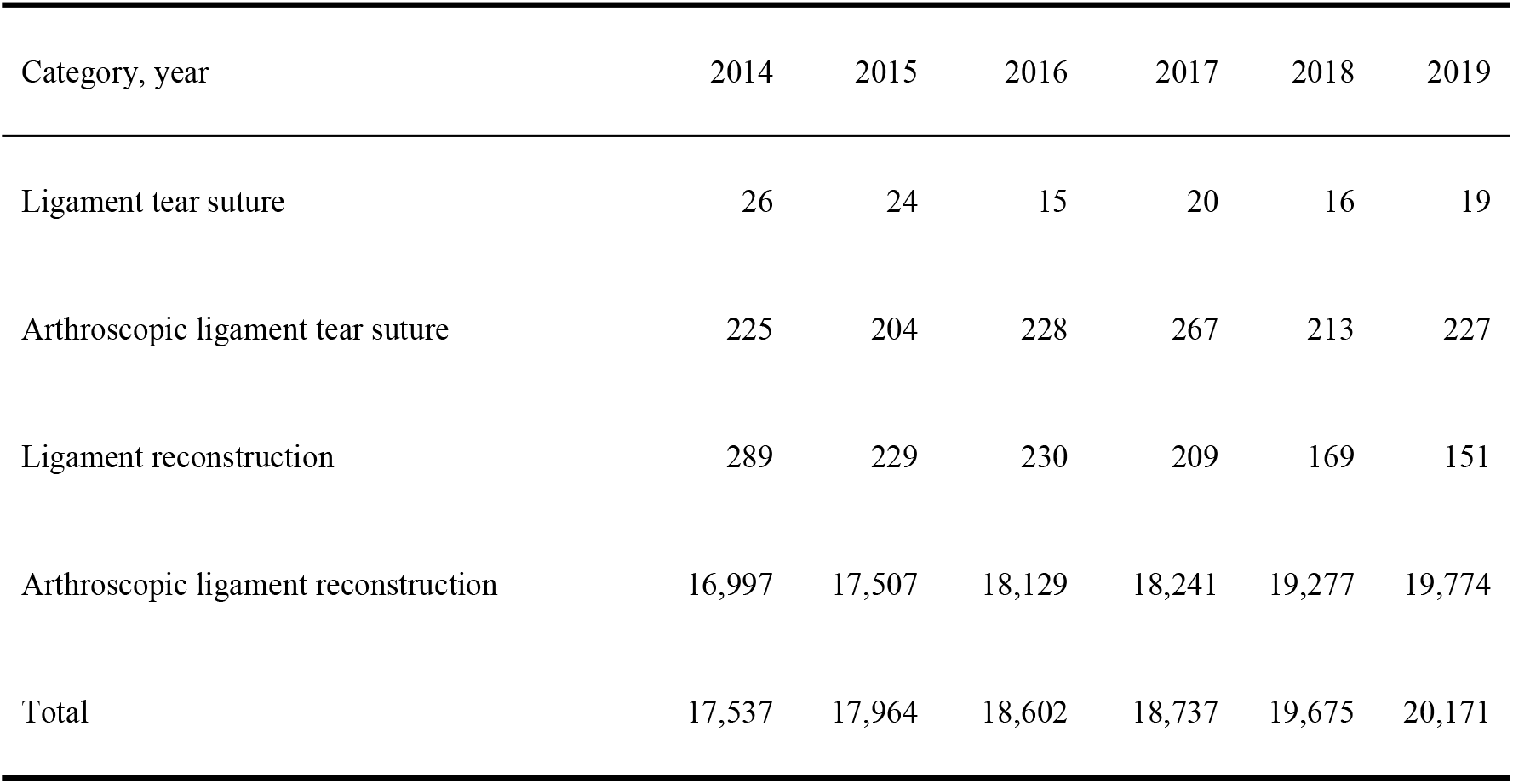
Annual number of cruciate ligament surgeries in the NDB open data from 2014 to 2019.

| Category, year | 2014 | 2015 | 2016 | 2017 | 2018 | 2019 |
| --- | --- | --- | --- | --- | --- | --- |
| Ligament tear suture | 26 | 24 | 15 | 20 | 16 | 19 |
| Arthroscopic ligament tear suture | 225 | 204 | 228 | 267 | 213 | 227 |
| Ligament reconstruction | 289 | 229 | 230 | 209 | 169 | 151 |
| Arthroscopic ligament reconstruction | 16,997 | 17,507 | 18,129 | 18,241 | 19,277 | 19,774 |
| Total | 17,537 | 17,964 | 18,602 | 18,737 | 19,675 | 20,171 |

Due to low registration numbers, data for the 0–9 and >70 years age groups were incomplete. For the 10–69 years age group, the highest number of registrations was observed in the 15–19 years age group (108.5 cases/100,000 population), followed by the 20–24 years age group (41.8 cases/100,000 population). We noted that registrations declined with age (Fig 2 and S2 and S3 Tables).

**Fig 2.**
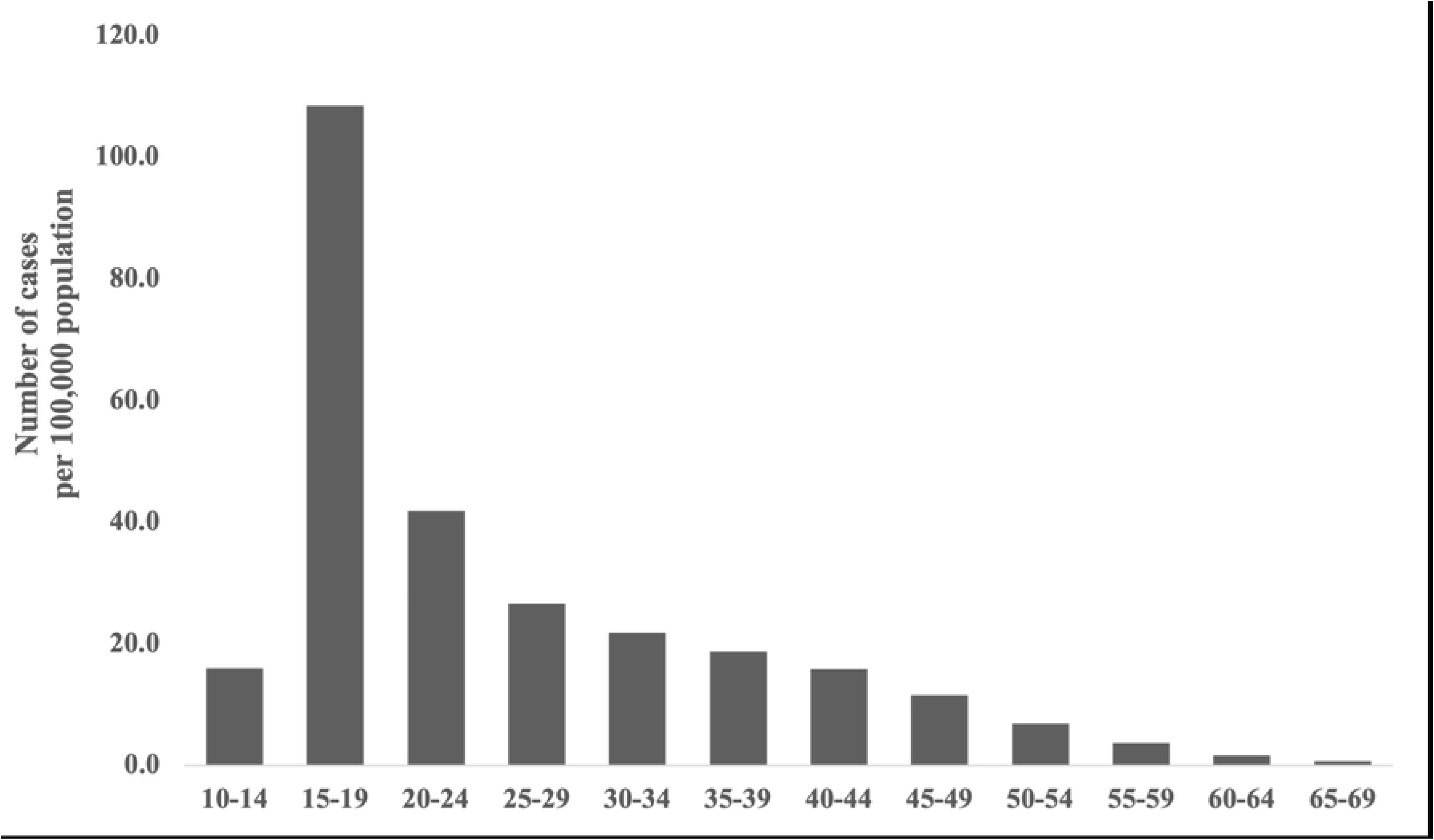
Number of registered arthroscopic ligament t reconstruction procedures per 100,000 population by age group from 2014 to 2019.

In terms of sex, females outnumbered males by a factor of 6.7 in the 10–14 years age group and by 1.6 in the 15–19 years age group. In contrast, males were 1.6 times more likely than females to be enrolled in the 20–24 years age group and more than twice as likely as females to be enrolled in the 25–29 and 30–34 years age groups. Registration numbers were similar for other age groups (Fig 3 and S3 Table).

**Fig 3.**
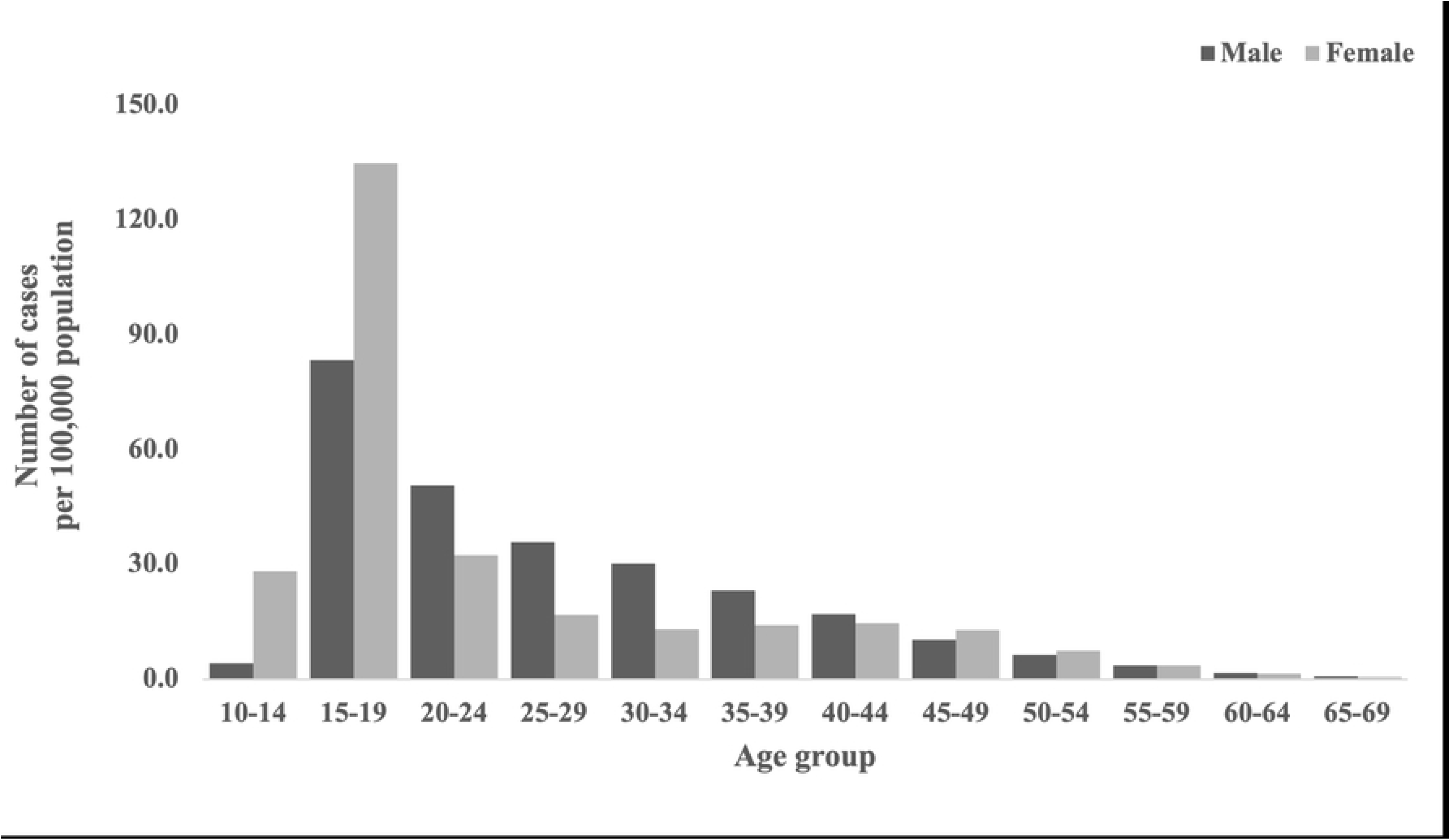
Number of registered arthroscopic ligament reconstruction procedures per 100,000 population.

## Discussion

This study investigated the number of registered CL surgeries, sex differences, and age characteristics in Japan from 2014 to 2019 using the NDB open data. As hypothesized, there was a consistent increase in CL surgery over the study period. Notably, most CL surgeries were arthroscopic ligament reconstruction procedures, which constitutes a significant finding of this study. With the advancement of medical technology, arthroscopic surgery, which enables a detailed examination of the interior of a joint during surgery, has become increasingly prevalent [21–23]. This development may have led to the rise in the number of arthroscopic procedures in Japan.

Arthroscopic ligament reconstruction was found to vary by sex and age. Overall, the incidence was higher among males. Conversely, females often undergo surgery during their teenage years, potentially due to the disparity in growth rates between males and females. Height growth in Japanese females reaches its peak around 2 years earlier than in males [24]. Variations in growth rates may influence the timing of surgery. However, it should be noted that CL surgery has been a subject of intense debate regarding its potential impact on growth retardation [25,26]. Other factors, such as female’s inability to be active in sports at times due to childbirth, may be related to the sex difference. Another finding was that the incidence of surgery among middle-aged and older adults had also increased. Injuries resulting from sports activities, everyday sprains, and falls are common among middle-aged and older adults [27]. This can be attributed to increased health awareness and higher physical activity levels. Additionally, there is a higher risk of knee joint dysfunction and secondary injuries (such as meniscus and cartilage damage) [28,29]. Good outcomes of CL surgery in middle-aged and older patients have also been reported [30,31]. Consequently, many patients may choose surgery with the goal of regaining their previous level of activity. In Japan, like in other countries, the indications for surgery have likely expanded due to changes in individual lifestyles and advancements in medical technology. This study had several limitations. First, it failed to provide details on the number of ACL and PCL surgeries as a categorization of the CL surgeries. Second, tissue injuries other than those caused by CL surgery were not identified, as they cannot be used for ligament and meniscus surgeries for health claims. Third, it was impossible to analyze information other than medical practice, sex, and age. Finally, traffic accidents and occupational injuries were not included in the present data.

In future studies, we aim to investigate the number of injuries and reconstruction surgeries separately for ACL and PCL injuries. We would also like to clarify the characteristics of Japanese patients through comparison with overseas reports. We believe organizing information on knee joint ligament injuries will lead to novel suggestions for rehabilitation and prevention programs.

## Conclusion

The present study identifies an increased incidence of CL surgery in Japan, along with gender and age characteristics. This trend is not limited to younger age groups but is also observed in middle-aged and older age groups. In the future, more detailed data should be extracted from medical databases to identify insights for optimal treatment strategies for specific populations.

## Data Availability

We utilized the NDB Open Data managed by the Ministry of Health, Labour and Welfare. Access to the dataset can be obtained through the website of the Ministry of Health, Labour and Welfare at [https://www.mhlw.go.jp/stf/seisakunitsuite/bunya/0000177182.html].

## Acknowledgments

This work was supported by JSPS KAKENHI Grant Numbers JP21K02905. We would like to express our gratitude to all the members of Taguri’s laboratory for their valuable discussions and comments. Additionally, we would like to thank Editage (https://www.editage.jp/) for providing English language editing services.

## Supporting information

S1 Table. STROBE Statement—checklist of items that should be included in reports of observational studies.

S2 Table. Annual registrations of arthroscopic ligament reconstruction according to age groups from 2014 to 2019. To avoid the identification of individuals, aggregate units that are <10 in principle are not included.

S3 Table. Annual registrations of arthroscopic ligament reconstruction per 100,000 population according to sex and age groups from 2014 to 2019. To avoid the identification of individuals, aggregate units that are <10 in principle are not included.

